# TB disease yield from household contact screening of tuberculosis index patients; a cohort study from Karachi, Pakistan

**DOI:** 10.1101/2023.04.17.23288669

**Authors:** Maria R. Jaswal, Saira Farooq, Hamidah Hussain, Jinsar Shah, Kumail Nasir, Ahsan Khalil, Hiba Khan, Nauman Safdar, Amyn A. Malik, Christopher F. McQuaid

**Affiliations:** Interactive Research and Development Global, Karachi, Pakistan; Global Health Directorate, Indus Hospital & Health Network, Karachi, Pakistan; TB Modelling Group, TB Centre and Centre for Mathematical Modelling of Infectious Diseases, Department of Infectious Disease Epidemiology, Faculty of Epidemiology and Population Health, London School of Hygiene & Tropical Medicine, London, UK

## Abstract

Nearly 40% of people affected by TB in Pakistan are not diagnosed each year. Guidelines recommend screening household contacts however, not all index patients or contacts are eligible. Therefore, many contacts who may have TB disease, remain unscreened.

We conducted a prospective cohort study under programmatic conditions in Karachi, Pakistan from January 2018 - December 2019, to screen all household contacts of all TB index patients. We disaggregated these according to guidelines into eligible (those with bacteriologically confirmed pulmonary TB or children <5 years) or ineligible (those with clinically diagnosed or extrapulmonary TB ≥5 years) index patients, and eligible (children <5 years or symptomatic individuals) or ineligible (asymptomatic individuals ≥5 years) contacts. We calculated TB disease yields for different groups of index patients and contacts.

Out of 39,168 household contacts from 6,362 index patients, 21,035 completed clinical assessments for TB disease, and 416 were diagnosed with all forms TB. Household contacts of clinically diagnosed pulmonary TB patients were 26% more likely to be diagnosed with TB compared to the household contacts of bacteriologically confirmed pulmonary TB (adjusted Odds Ratio 1.26 [1.01 – 1.59] p-value:0.03). The yield of TB disease among child contacts (3.4%) was significantly higher than the yield among adult contacts (0.5%) (*p*-value:<0.001).

Broadening TB contact screening guidelines to include clinically diagnosed and extrapulmonary index patients ≥5 years could double the number of patients detected at a similar level of effort.

## Introduction

About 10 million people worldwide fall sick with tuberculosis (TB) annually, around 2.9 million of whom are never diagnosed or reported (*1*). In Pakistan, the rate of underdiagnoses increases to an estimated 220,000 of 570,000 TB cases (39%), with approximately 46,000 people dying of TB annually. A 2018 United Nations High-Level Meeting on the fight against TB agreed to targets for diagnosing and treating missing TB patients (*2*), and the World Health Organization (WHO) has since released guidelines for countries to implement the End TB Strategy at a national scale to achieve these targets (*3, 4*). These guidelines recommend various active case finding strategies, one of which is TB contact screening. This consists of evaluating household contacts of index patients for TB disease, as these individuals may be at an increased risk of TB disease themselves. Pakistan’s 2019 National TB Report (*5*) recorded ∼200,000 household contacts screened in 2018.

Pakistan’s National TB Control Program (NTP) guidelines are in line with the older WHO recommendations (*6*) for contact screening. These categorise TB patients as follows: pulmonary tuberculosis with bacteriological confirmation; pulmonary tuberculosis established clinically; and extrapulmonary tuberculosis (*4*). TB index patients eligible for household contact screening according to these guidelines are those diagnosed with pulmonary tuberculosis with bacteriological confirmation (considered infectious) and children <5 years diagnosed with TB (to identify the source case) (*6*). TB patients ≥5 years old who are clinically diagnosed with pulmonary TB or are diagnosed with extrapulmonary TB are not eligible for contact screening (Figure-1).

**Figure 1:**
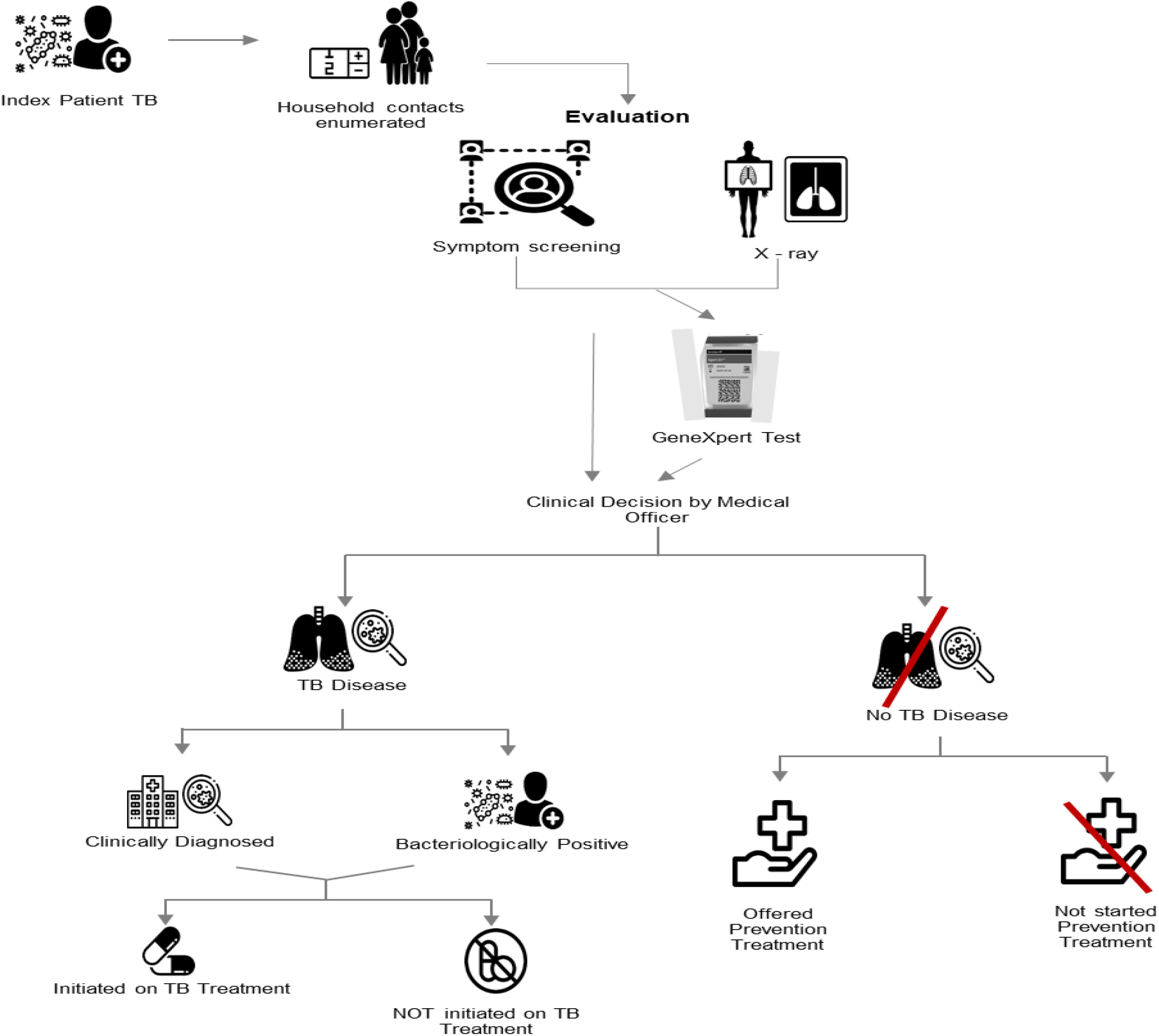
Patient flow from the initial diagnosis of a TB index patient to complete screening and evaluation of household contacts for TB diagnosis and prevention treatment.

Household contacts of TB patients are divided into the following 3 categories: children <5years old; individuals ≥5 years old with TB symptoms; and individuals ≥5years old without symptoms (asymptomatic). Children <5years old and individuals ≥5 years old with TB symptoms who are household contacts of eligible index patients (either TB patients with bacteriologically confirmed TB or TB patients <5years old) are eligible for contact screening (*7*). Asymptomatic contacts are not eligible for TB screening. Guidelines defining eligible index TB patients and contacts are elaborated in figure 1.

However, more recent WHO guidelines recommend contact screening of all close contacts of all types of TB index patients (*4*), if resources permit. Due to lack of resources, Pakistan’s guidelines remain unchanged, potentially resulting in missed TB patients among household contacts of ineligible TB index patients. Several studies have reported TB case detection among contacts of clinically diagnosed pulmonary TB and extra-pulmonary TB patients (*8, 9*). Extending contact screening guidelines to include all index patients and all their household contacts could help in the identification of additional persons with TB, and the prevention of further spread of infection. Here, we estimate the frequency of TB cases among individuals eligible and ineligible for screening according to current Pakistan guidelines, in a prospective cohort of index patients under programmatic conditions in Karachi, Pakistan.

## Materials and Methods

### Study Design and Participants

A prospective cohort study was conducted in Karachi, Pakistan, screening all household contacts of all types (including all of pulmonary TB and extrapulmonary TB) of drug-susceptible TB index patients enrolled at eight TB clinics between January 2018 and December 2019. We used a previously defined criteria for household contact with some additional details. Household contact was defined as a person sharing the same kitchen under the same roof with an average exposure to a TB patient of 6-8 hours a day for 5-7 days a week during the last 3 months (*10*). A household contact was excluded from the study if s/he was receiving TB treatment or TB preventive treatment at the time of the first contact screening interview.

The study was approved by the Institutional Review Board of Interactive Research and Development and the National Bioethics Committee of Pakistan.

### Study Procedures

All TB patients at the time of enrolment were counselled by the study counsellor on the importance of contact screening and TB preventive treatment. The invitation and consent process has been outlined elsewhere (*11*).

All household contacts who consented/assented underwent a chest x-ray, after which they were seen by the medical officer from the program. The medical officers were trained in the assessment and diagnosis for TB, especially for child TB diagnosis and were additionally supervised by adult and child TB consultants. The medical officer conducted a thorough verbal symptoms screen along with medical history, physical examination (where needed) and assessment of chest x-ray. Contacts were recorded as symptomatic if they had any of the following TB symptoms present; cough, fever, weight-loss or night sweats, as defined by the NTP guidelines (*6*). Clinical evaluation included additional questions for the assessment of extrapulmonary TB. Complete TB disease evaluation was defined as having a chest x-ray, sputum examination (if able to produce sputum) and disease evaluation with medical officer which included complete medical history, physical examination and any further lab investigations requested by the medical officer for the diagnosis of TB disease. For children <5 years, mid upper arm circumference (MUAC) was recorded to assess the nutritional status (*12*). For all children 0-14years, CDC growth charts for weight-for-age percentiles were used to assess the nutritional status (*13*). For adults ≥15 years, body mass index (BMI) was considered for nutritional status.

If the contact had TB symptoms and/or x-ray suggestive of TB, or if they were able to produce sputum, they were asked to submit a sputum sample for Xpert MTB/RIF testing (Cepheid, Sunnyvale, CA, USA). In cases where the contact was unable to produce sputum, the medical officer diagnosed/ruled out TB disease on clinical grounds. Wherever needed, the medical officer requested further lab investigations to reach the diagnosis, including complete blood count, Erythrocyte sedimentation rate, C-reactive protein, tuberculin skin test, ultrasound, computer tomography scan, fine needle aspirate cytology, and histopathology of body tissue sample. In the absence of bacteriological confirmation, TB diagnosis was made as per the discretion of the medical officer. All contacts diagnosed with TB disease were linked to TB disease treatment on-site, and contacts without TB disease were offered TB preventive treatment by the program.

### Outcome Measure & Statistical Analysis

#### Index patients

Eligible index patients according to NTP guidelines include:

○ <5 years diagnosed with TB (all types)
○ ≥5 years diagnosed with bacteriologically confirmed pulmonary TB

Ineligible index patients include:

○ ≥5 years diagnosed with pulmonary TB established clinically
○ ≥5 years diagnosed with extrapulmonary TB

#### Household contacts

Eligible household contacts according to NTP guidelines include:

○ <5years old household contacts of eligible index TB patients
○ ≥5years old household contacts of eligible index TB patients with TB symptoms

Ineligible household contacts include:

○ <5years old household contacts of ineligible index TB patients
○ ≥5years old household contacts of ineligible index TB patients with TB symptoms
○ ≥5years old household contacts of eligible or ineligible index TB patients without TB symptoms

#### Outcome measure

The outcome measure was “yield of TB disease”, defined as the percentage of people with TB diagnosed out of the total number evaluated. Yield of TB disease was calculated for each group of household contacts disaggregated by age group and by index patient type. We also recorded the number of contacts needed to screen to find one contact with TB disease (NNS), calculated as follows:

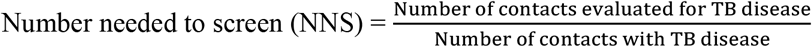

Difference in proportions was calculated using chi-squared analysis and we considered there to be statistically significant difference in proportions if the p-value was <0.05. The odds ratio (OR) was calculated to compare the odds of TB disease among the contacts of eligible index TB patients to the odds of TB disease among contacts of ineligible index TB patients.

## Results

From January 2018 to December 2019, a total of 6,361 TB index patients were enrolled at the eight TB treatment sites in Karachi (Supplementary table 1). There were a total of 39,168 contacts from all index patients with an average of 6.2 household contacts per index patient and majority of the contacts were adults (57.2%). However, nearly two thirds (63.9%) of child contacts completed clinical evaluation for TB disease compared to less than half of adult contacts (46.1%). Complete TB disease evaluation differed significantly between contacts of eligible and ineligible TB index patients, with a higher proportion (56.6%) seen among contacts of ineligible TB index patients compared to the proportion (51.3%) among contacts of eligible TB index patients (*p*-value:<0.001).

A total of 416 contacts were diagnosed with TB (see Table 1), 50.8% (n=211) of whom were female (Table 2) and 56.5% (n=227) underweight compared to the standard for their age (14, 15). Cough, fever and weight-loss were the most commonly reported symptoms. Most of the contacts diagnosed with TB disease were household contacts of child TB index patients (58.8%). The yield of TB disease among child contacts <15years (3.4%) was significantly higher than the yield among adult contacts ≥15years (0.5%) (*p*-value:<0.001)

**Table 1:**
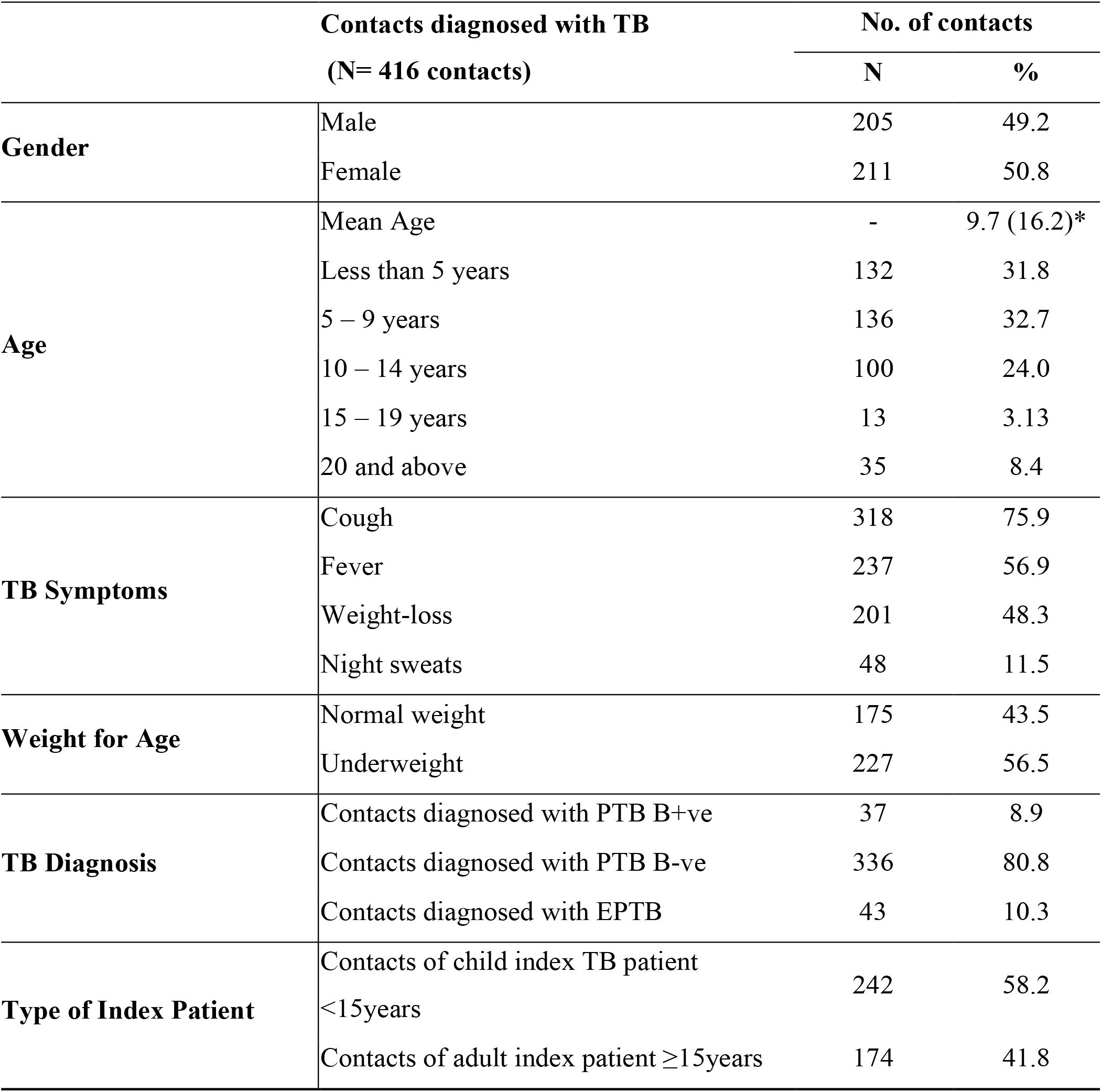
Demographics and characteristics of contacts diagnosed with TB from all forms TB index patients. PTB=pulmonary TB, B+ve=bacteriologically confirmed, B-ve=clinically diagnosed, EPTB=extra-pulmonary TB. *standard deviation calculated for the mean age

**Table 2:** Odds ratios and stratified analysis for contacts diagnosed with TB from eligible and ineligible index patients.

|  | Odds ratio | (95% confidence interval) | p-value |
| --- | --- | --- | --- |
| Yield from ineligible index patients with eligible index patients as baseline group | 0.99 | 0.82 – 1.20 | 0.95 |
| **Stratified Analysis** |  |  |  |
| **Index patient type** |  |  |  |
| Bacteriologically confirmed pulmonary TB $\geq 5$ years | base | - | - |
| Clinically diagnosed pulmonary TB $\geq 5$ years | 1.26 | 1.01 – 1.59 | 0.04 |
| Extrapulmonary TB $\geq 5$ years | 0.86 | 0.62 – 1.19 | 0.37 |
| **Index age group** |  |  |  |
| <5 years | base | - | - |
| $\geq 5$ years | 0.70 | 0.54 – 0.92 | 0.01 |
| **Contact Gender** |  |  |  |
| Female | base | - | - |
| Male | 0.93 | 0.76 – 1.14 | 0.51 |
| **Contact Age** |  |  |  |
| <5 years | base | - | - |
| 5-9 years | 0.90 | 0.70 – 1.15 | 0.42 |
| 10-14 years | 0.85 | 0.65 – 1.11 | 0.24 |
| 15-19 years | 0.16 | 0.09 – 0.29 | <0.001 |
| $\geq 20$ years | 0.11 | 0.07 – 0.16 | <0.001 |

A total of 191 TB patients were diagnosed when guideline-defined eligible household contacts from eligible TB index patients were evaluated for TB disease. Through the screening of ineligible household contacts, 225 contacts were diagnosed with TB disease (Figure 2). TB disease yield was found to be similar for household contacts <5years and symptomatic contacts ≥5 years from guideline-eligible (6.3%) and ineligible (6.0%) TB index patients (*p*-value:<0.71). TB disease yield among asymptomatic household contacts from guideline-defined eligible and ineligible TB index patients was significantly lower, at 0.3-0.4%.

**Figure 2:**
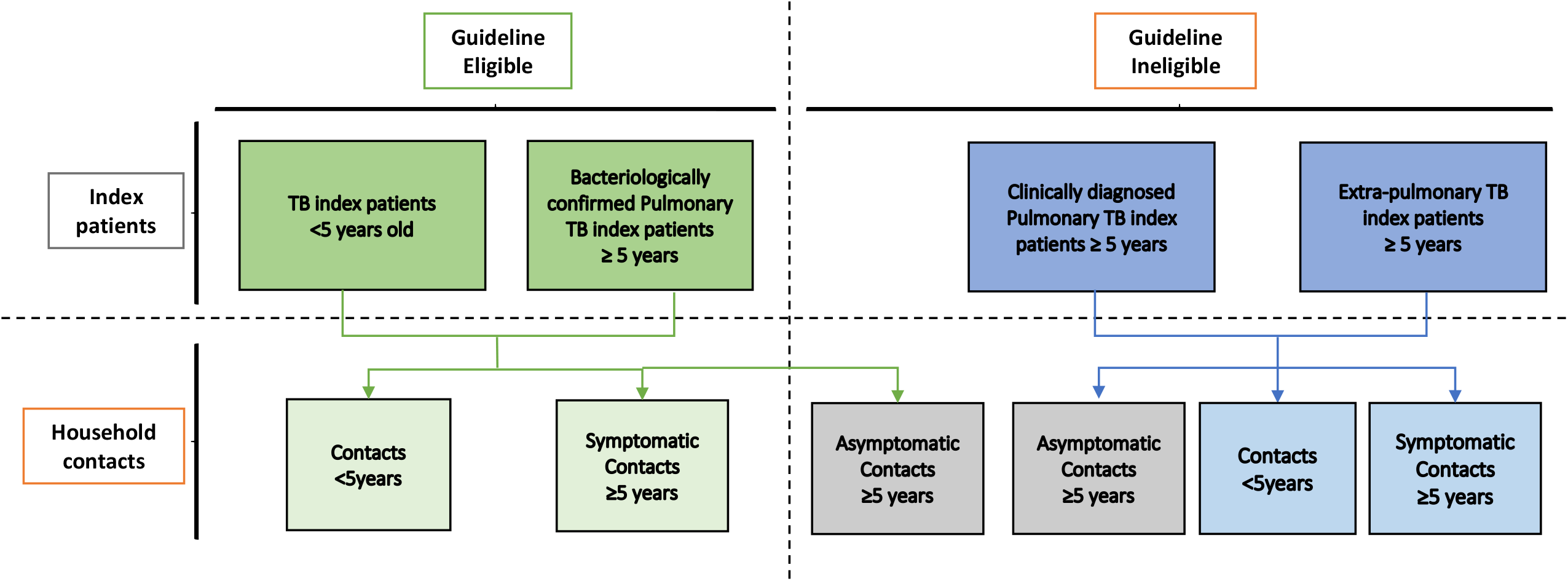
TB index patients and their household contacts segregated into groups according to the Pakistan National TB Control Program (NTP) guidelines into eligible and ineligible categories

Odds of TB disease (Table 2) were similar among contacts of eligible and ineligible index TB patients (OR: 0.99 95% Confidence Interval [CI]: 0.81-1.20). There was strong evidence for higher odds of TB disease in contacts of a clinically diagnosed pulmonary TB index patient compared to an index patient with bacteriologically confirmed pulmonary TB, after adjusting for age of index patients and age and gender of the contacts (adjusted OR [aOR]: 1.26, 95% CI: 1.01 – 1.59). However, the adjusted model showed no association of TB disease among contacts of extra-pulmonary TB index patients ≥5 years compared to bacteriologically confirmed pulmonary TB (aOR: 0.86, 95% CI: 0.62 – 1.19).

The number needed to screen for symptomatic contacts ≥5 years was 10 in the guideline-defined eligible and ineligible TB index patients (Figure-3). Among child contacts <5years old, NNS was 25 in guideline-eligible index TB patients and 29 in guideline-ineligible index TB patients. However, NNS was higher among asymptomatic contacts, ranging from 244-314 for guideline-defined eligible and ineligible index TB patients respectively.

**Figure 3:**
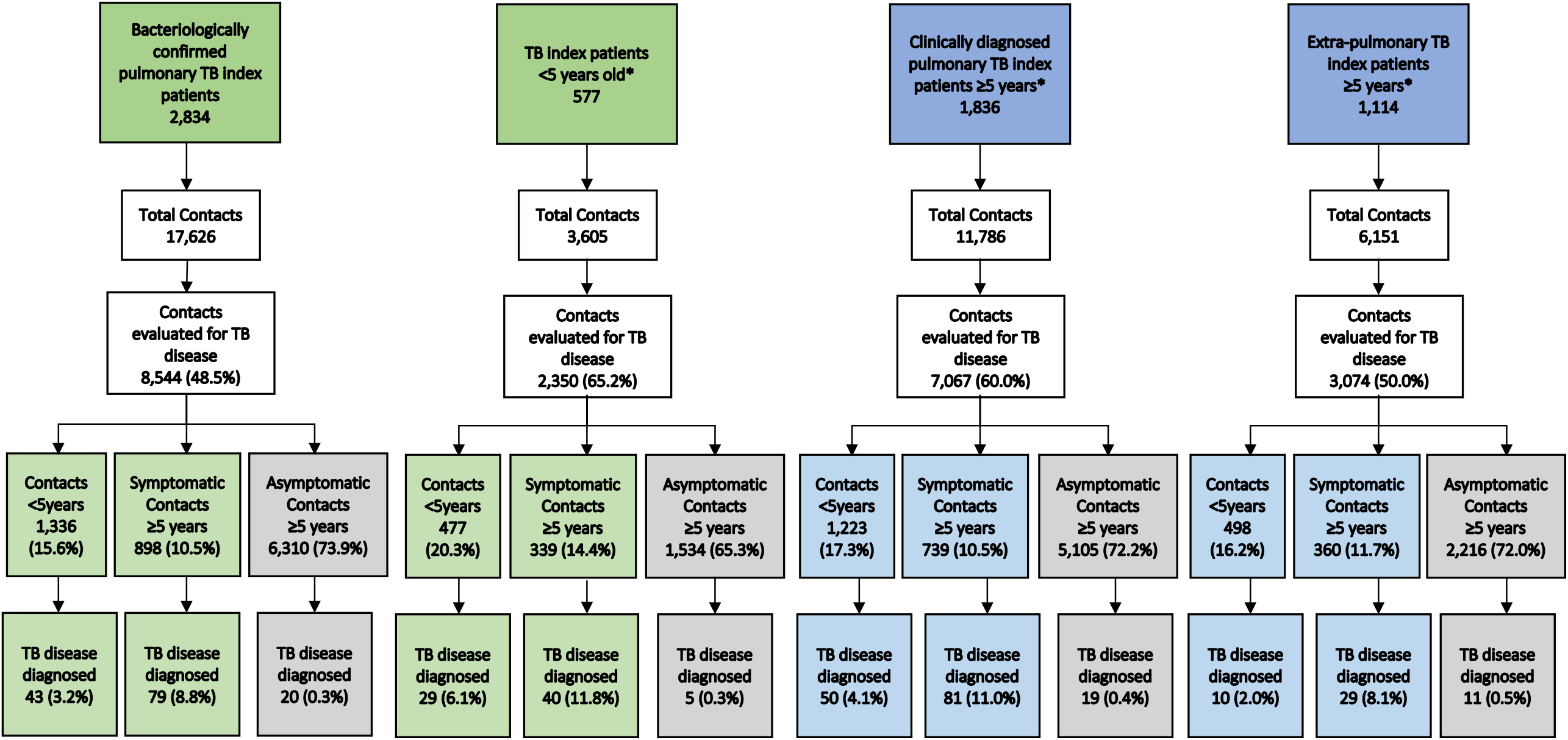
Flow diagram showing index patients, household contacts and TB disease yields for individuals eligible for household contact screening according to Pakistan’s National TB Control Program (NTP) Guidelines compared to TB index patients and household contacts not eligible for contact screening. Light green and dark green boxes represent TB index patients and their household contacts eligible for household contact screening as per NTP guidelines. Dark blue boxes represent TB index patients ineligible for household contact screening as per NTP guidelines, and their household contacts (light blue boxes) that would be eligible if the index patient eligibility criteria were extended. Grey boxes represent household contacts that would be eligible if the contact eligibility criteria were extended, for eligible and ineligible index TB patients respectively. **\***Excluding bacteriologically confirmed pulmonary index TB patients

**Figure-4:**
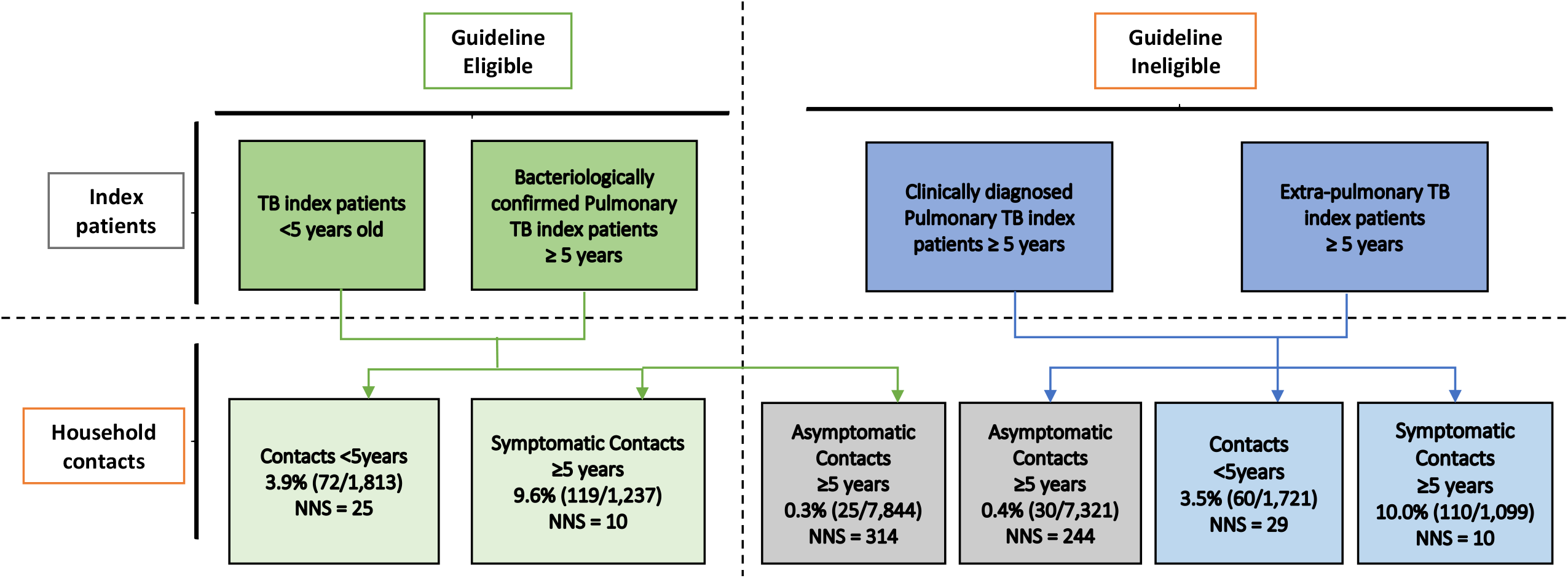
TB index patients and their household contacts segregated into groups according to the Pakistan National TB Control Program (NTP) guidelines into eligible and ineligible. TB disease yield is shown in each group of contacts. Light green box represents the guideline eligible contacts and their disease yield. Light blue boxes represent the group of ineligible contacts that is recommended for inclusion in guidelines for contact screening.

## Discussion

In the first study conducted in Pakistan to evaluate TB disease yields among household contacts of all types of TB index patients, we found the risk of TB in certain household contacts (<5 years and symptomatic contacts) is as high in TB index patients ineligible for contact screening as it is for TB index patients eligible for contact screening. TB disease yield from contacts of TB index patients ineligible for screening under current guidelines (1.97%) was similar to the yield (1.98%) from eligible index patients (OR: 0.99 [0.82 – 1.20]). Additionally, household contacts of clinically diagnosed pulmonary TB patients were 26% more likely to be diagnosed with TB compared to the household contacts of bacteriologically confirmed pulmonary TB patients after adjusting for age and gender of the contacts. Expanding the index patient eligibility criteria to include high-risk contacts of clinically diagnosed pulmonary TB and extrapulmonary TB patients would have resulted in an additional 170 TB diagnoses, compared to 191 under the current guidelines (170+191=361; ∼90% increase). Implementation of this study under programmatic conditions suggests that this could significantly improve rates of diagnosis for important groups at risk of TB disease.

Current NTP guidelines in Pakistan emphasise household contact screening for the index TB patients with bacteriologically confirmed pulmonary TB, as it is considered to be the most infectious form of TB (*6*). However, our study found 150 TB patients from household contact screening of clinically diagnosed pulmonary TB index patients, emphasizing the importance of screening in this group of contacts. Meanwhile, extrapulmonary TB patients are considered to be non-infectious; however, screening contacts of extrapulmonary TB patients is known to yield disease (*8, 16*). Our analysis shows that, after adjusting for age and gender of contacts and age of index patients, household contacts of extrapulmonary TB index patients are less likely to be diagnosed with TB disease than contacts of bacteriologically confirmed pulmonary TB index patients. However, we diagnosed 50 additional contacts with TB from extrapulmonary TB index patients. Therefore, screening in contacts of clinically diagnosed pulmonary TB and extrapulmonary TB index patients would find missing TB patients that are essential to realise TB targets (*17*).

Recent guidelines recommend screening for contacts <5 years and those with TB symptoms (*4, 6*). When conducting screening for this group of contacts from guideline-defined eligible index TB patients, we found 191 contacts with TB disease. When this high-risk group of contacts from guideline-defined ineligible index TB patients was screened, we found 170 contacts with TB disease. NNS to find one TB patient in this group of contacts from eligible index TB patients was 16 compared to NNS of 17 in the same group of contacts from ineligible index TB patients. However, screening asymptomatic contacts of all index patients had a significantly lower, but non-zero, yield (0.36%) than screening the high-risk group of contacts (6.15%). The large number of contacts in this category means that this still represents a significant proportion (here∼13%) of total people with TB (*18*). Indeed, one South African study reported a majority of contacts diagnosed with TB to be asymptomatic (*19*). These individuals, if left undiagnosed and untreated, could be a source of continued transmission in the community (*20*). Screening these contacts also provides the opportunity to start many on TB prevention treatment, which requires TB evaluation to rule out TB disease before treatment initiation (*7*).

We also found a higher TB yield among contacts of child TB index patients than adults, similar to the studies from Southern Africa (*21*). Children are frequently exposed to TB at home through adult caregivers (*22*), where contact screening serves to identify both the source and other exposed children (*23, 24*). It is of importance to note that the index patient is not necessarily the source case (*25*), but rather the one on whom the contact screening is centred. Restricting index patient criteria to infectious TB patients only may miss opportunities for diagnosing the actual source case.

Overall, significantly higher TB yields were observed among child contacts from all forms TB index patients, where the yield remained higher among this age group when segregated by the type of TB in the index patient or the age of the index TB patient (*22, 24*). Among all contacts diagnosed with TB, more than 85% were <15 years and ∼32% were <5 years. The latter are considered to be at high risk of developing TB infection and TB disease (*26*) and are already prioritized in WHO guidelines (*4*), however the former are currently excluded, where our results support previous studies that have found a high yield of TB disease in this group (*21*).

Among enumerated contacts, we found more adults than children. However, a significantly higher proportion of children completed the disease evaluation process (64%) compared to adults (46%), similar to other studies in Pakistan (*24*), which could affect our results. Low evaluation rates, particular among adults, have been observed as a result of an inability to visit the facility due to work, financial or transport constraints, or when contacts feel healthy or are concerned about stigma (*23, 27*). As the recent TB prevention treatment guidelines recommend TB prevention treatment for household contacts of all ages (*28*), efforts should be increased to improve disease evaluation among contacts of all ages.

Our results are like many previous studies from Asia and Africa of all pulmonary index TB patients (*26, 27, 29*). Studies from Pakistan and elsewhere have included only bacteriologically confirmed pulmonary index TB patients and screened only symptomatic contacts or those <5 years, reporting comparable or higher yields (*30, 31, 32, 33*). One study from Pakistan has reported a significantly higher TB yield of ∼15% among household contacts of child TB index patients (*23*). Other studies with higher yields (19, 34) do not allow for a comprehensive comparison of different forms of TB index patients and their contacts. We have also found more child contacts with TB than adult contacts, similar to studies from Pakistan (*24*) and India (*22*).

One of the limitations in our study is the possibility of over-diagnosis in the clinically diagnosed TB patients, although in high TB burden settings a proportion of false positive TB cases among key and vulnerable populations may be acceptable since the benefit of treatment is still higher than no treatment at all (*35*). We did not test for HIV infection as it is not routinely done in our context given the low HIV incidence. Another limitation of the study was inability to evaluate all the enumerated household contacts which may have resulted in over or under estimation of TB disease yield. However, proportion of contacts evaluated across eligible and ineligible groups is similar, therefore, the difference in yield is not expected to change. Additionally, our study was conducted in one of the major metropolitan cities in the country, and results may not be generalizable to rural settings or peri-urban areas with differing household structures and practices (*19*). However, other studies have also found similar results in rural settings (*24*).

Overall, our results suggest that a significant proportion of TB in household contacts can be found in contacts who are currently ineligible for screening, either due to index patient or contact ineligibility. Broadening TB contact screening guidelines to include clinically diagnosed and extrapulmonary index patients ≥5 years could double the number of TB patients detected at a similar level of effort. However, efforts to end TB should also consider the role and cost-effectiveness of screening asymptomatic household contacts ≥5 years.

## Data Availability

The de-identified dataset analyzed for the current study is available from the corresponding author on reasonable request and on signing of a data-sharing agreement

## Authors’ contribution

MRJ, AAM conceptualized the study and wrote the protocol; SF, KN, AK, and JS collected data under supervision from NS, and HH; SF, HK and MJ had access to primary data and verified the dataset; AAM, SF, MJ, performed and reviewed the analysis; MJ, CFM, AAM and HH wrote the initial draft of the manuscript. All authors helped interpret the findings, read, and approved the final version of the manuscript.

## Acknowledgements

We would like to acknowledge the participants and site management staff, all the program team. Additionally, we would like to thank Pauline Scheelback for her input and some insight about structuring the argument.

## Author’s biographical sketch

Author is an early-career global health researcher based in Karachi. Her experience is with TB program implementation including TB clinic, TB active case finding and prevention treatment. Her interest lies in disease prevention and cost-effectiveness of programs at scale.

