## Supplementary figures and images for "TB disease yield from household contact screening of tuberculosis index patients; a cohort study from Karachi, Pakistan"

### Supplemental Table

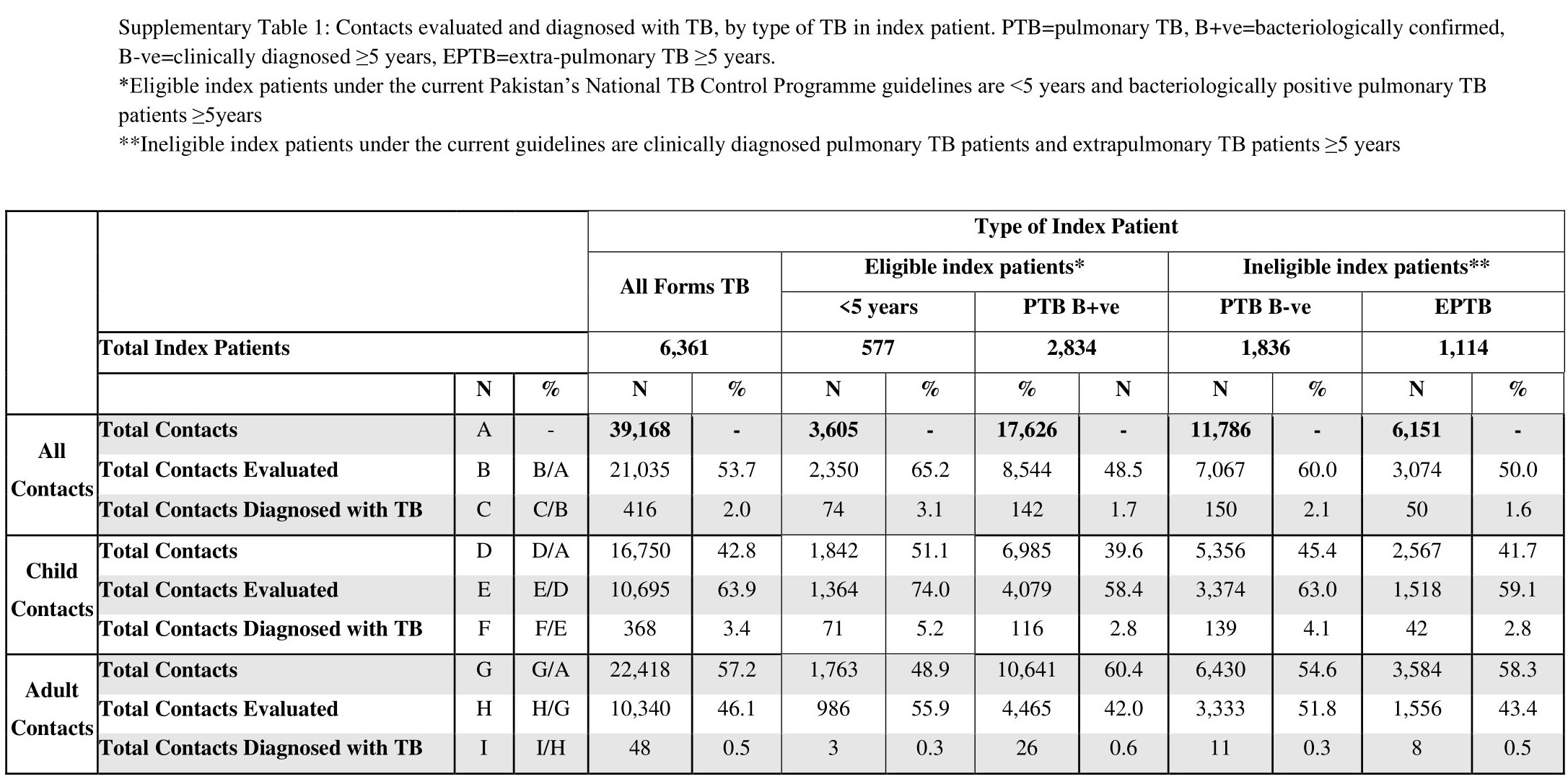
